# Safety of Pharmacologic Dilation: Acute Angle Closure Incidence in a Los Angeles County-Wide Safety Net Teleretinal Screening Program

**DOI:** 10.1101/2025.06.26.25330091

**Authors:** Tracy Z. Lang, Benjamin Y. Xu, Zhiwei Li, Sreenidhi Iyengar, Carl Kesselman, Jose-Luis Ambite, Kyle Bolo, Jiun Do, Brandon Wong, Lauren P. Daskivich

**Affiliations:** Roski Eye Institute, Keck School of Medicine, University of Southern California, Los Angeles, CA, USA; Information Sciences Institute, Viterbi School of Engineering, University of Southern California, Los Angeles, CA, USA; Los Angeles General Medical Center, Department of Ophthalmology, Los Angeles County, Los Angeles, CA, USA; Ophthalmology and Eye Health Programs, Department of Health Services, Los Angeles County, Los Angeles, CA, USA

**Author notes:** **Corresponding Author:** Lauren P. Daskivich, MD, MSHS, Ophthalmology and Eye Health Programs, Department of Health Services, Los Angeles County, 313 N Figueroa St, Ste 904C, Los Angeles, CA 90012. Tracy Z. Lang and Benjamin Y. Xu contributed equally as co-first authors.

## Abstract

**Importance:** Pharmacologic dilation is vital for eye disease screening but is often avoided due to concerns about triggering acute angle closure (AAC), a sight-threatening ophthalmic emergency.

**Objective:** To assess AAC incidence after dilation and validate the use of International Classification of Diseases (ICD) codes for identifying AAC cases.

**Design:** Retrospective cohort study

**Setting:** Primary care-based teleretinal diabetic retinopathy screening (TDRS) program

**Participants:** Eligible participants were Los Angeles County (LAC) Department of Health Services (DHS) patients who underwent teleretinal screening by dilated fundus photography between August 23, 2013, and March 1, 2024. Potential AAC cases were identified using ICD codes for angle closure, including acute angle closure glaucoma (AACG), primary angle closure glaucoma (PACG), and anatomical narrow angle (ANA), within three months of dilation. All urgent care, emergency department, and eye clinic encounters within the next calendar day after TDRS and encounters with Current Procedural Terminology (CPT) codes for iridectomy/iridotomy or lens extraction within 14 calendar days of TDRS were also identified. Manual chart review was conducted to verify AAC cases and extract clinical information.

**Exposures:** Dilation with 1.0% or 0.5% tropicamide.

**Main Outcomes and Measures:** Cumulative incidence of AAC after dilation.

**Results:** 84,008 patients received 168,796 dilations with a mean of 2.01 ± 1.50 (mean ± standard deviation) dilations per patient. 55.1% were female. Mean age was 55.4 ± 10.7 (mean ± standard deviation) years. The cohort was 67.7% Hispanic, 8.2% Black, 6.3% Asian, 4.1% White, and 2.4% Other. Manual chart review confirmed four AAC cases after dilation: 3 coded as AACG and 1 as ANA. The AAC risk was 2.4 (95% CI 0.05-4.69) per 100,000 dilations (0.0024%) or 4.8 (95% CI 0.10-9.43) per 100,000 patients (0.0048%). All four cases were female, had narrow angles in the non-presenting eye on gonioscopy, and presented within one day with AAC symptoms, including eye pain and blurry vision.

**Conclusions and Relevance:** AAC risk was less than 1 in 40,000 per dilation in a high-volume TDRS program serving a diverse, safety net population, supporting the overall safety of dilation in this setting. Further discussion about AAC risk as a contraindication to dilation is warranted.

## INTRODUCTION

Pharmacologic pupillary dilation is fundamental to comprehensive eye exams and improves detection of retinal and optic nerve pathology, including diabetic retinopathy (DR) and glaucomatous optic neuropathy. Dilation also enhances teleretinal screening by fundus photography, an alternative care delivery method that has increased in popularity as eye care demand rises and available workforce declines.^1,2^ Studies show that pupillary dilation reduces ungradable fundus photographs from 25-30% to 5-10% and increases the sensitivity for detection of DR compared to indirect ophthalmoscopy.^2-5^ Despite the clear diagnostic benefits of pharmacologic dilation, its use is not widely adopted by non-eye care providers primarily due to concerns about triggering an acute angle closure (AAC) attack.

AAC is an ophthalmic emergency that can lead to dramatic rise in intraocular pressure (IOP), rapid glaucomatous damage, and irreversible vision loss.^6-13^ Although previous studies consistently demonstrate very low AAC risk with dilation, ranging between 0.003% to 0.06% per dilation, concerns about AAC continue to deter its widespread use for eye disease screenings.^7-11^ These concerns may be related to the narrow demographics of previous studies, which primarily focused on White and Asian populations.^6-13^ In addition, many of these studies were cross-sectional and/or limited by small sample sizes. Therefore, there is a further need to evaluate the safety of pharmacologic dilation, particularly in high-volume screening settings serving diverse populations.

This study addresses these knowledge gaps by assessing AAC risk after pharmacologic dilation using data from the Los Angeles County (LAC) Department of Health Services (DHS) Teleretinal Diabetic Retinopathy Screening (TDRS) program. LAC DHS is the second largest municipal health system in the United States, serving over 700,000 primarily disadvantaged and uninsured patients annually. The TDRS program, launched in 2013, is a primary care-based program screening over 20,000 LAC DHS patients annually for DR and other eye diseases using mydriatic fundus photographs remotely reviewed by certified optometrist graders.^14-15^ The program increased DR screening rates by 16.3%, reduced wait times by 89.2% between 2013 to 2015, and detected manifest glaucoma after in-office evaluation in nearly a quarter of patients referred and evaluated for glaucoma between 2016 to 2018.^14,15^

Although the TDRS program has achieved a much lower rate of ungradable photos than screening programs that avoid pharmacologic dilation, the safety of this practice and incidence of AAC in the TDRS program are unknown.^2-4^

We also addressed the lack of evidence supporting the use of International Classification of Diseases (ICD) codes for identifying AAC cases. Although ICD codes have been validated for identifying cases of primary open angle glaucoma (POAG), this may not generalize to cases of AAC and primary angle closure glaucoma (PACG).^16-21^ Therefore, we validated a codified AAC phenotype through manual review of electronic healthcare records (EHR) to strengthen our study methodology and future big data studies on AAC.

## METHODS

### LAC DHS TDRS and Eligibility Criteria

TDRS patients underwent fundus photography at one of 17 LAC DHS teleretinal screening sites. Patients could walk in for same-day screening or have appointments scheduled in advance by their primary care physician or care manager. Eyes were dilated with one drop of 1.0% tropicamide ophthalmic solution at all sites throughout the study period, except for one site that used 0.5% tropicamide for several years due to facility-specific formulary restrictions. Eyes were dilated approximately 10 minutes prior to fundus photography unless good quality photos could be obtained without dilation, the patient declined, or the patient had a previous adverse reaction to dilation. Trained photographers obtained fundus photos according to a 3 45-degree standard-field protocol and uploaded the photos to a web-based screening software (EyePACS, EyePACS LLC, Santa Cruz, CA). Photos were graded by certified graders (LAC DHS optometrists) for DR, and findings consistent with other ocular pathology such as glaucoma, cataract, and age-related macular degeneration were also identified. Further details regarding program protocols have been described elsewhere.^14^ Patients were instructed to promptly visit the nearest Urgent Care Center (UCC) or Emergency Department (ED) if they experienced eye pain or discomfort within the next eight hours.

The study cohort included LAC DHS patients with diabetes who underwent teleretinal screening between August 23, 2013, and March 1, 2024. Eligible TDRS patients must not have received care from an eye care professional in the past 12 months. Exclusion criteria included missing identification numbers or no record of pharmacologic dilation during the screening visit. This study adhered to the tenets of the Declaration of Helsinki and was granted approval by the USC Institutional Review Board. This study follows the Strengthening the Reporting of Observational Studies in Epidemiology (STROBE) guidelines for cohort studies.

### Data Collection

TDRS records were cross-referenced with the LAC DHS electronic healthcare records (EHR) database. TDRS patients who received an ICD-9/10 diagnosis of acute angle closure glaucoma (AACG; 365.22, H40.21x), primary angle closure glaucoma (PACG; chronic: 365.23, H40.22*, intermittent: H40.23*, residual: H40.24*, unspecified: H40.20, H40.20X*), primary angle closure (PAC; 365.06, H40.06*), or anatomical narrow angle (ANA; 365.02, H40.03x) within three months of TDRS dilation were identified from all encounters in the LAC DHS system.

All UCC/ED and eye clinic encounters with eye-related diagnosis codes occurring within one calendar day of TDRS and all encounters with a Current Procedural Terminology (CPT) code for iridotomy, iridectomy, or lens extraction within 14 calendar days of dilation were also identified (See eTable 1 in the Supplement) to ensure that no AAC cases were miscoded and missed by our ICD code approach. Eye-related diagnosis codes included all ICD codes for ocular conditions, encompassing both primary and secondary eye conditions. Symptoms associated with acute angle closure, such as ocular pain (H57.1*), headache (R51.*), nausea (R11.*), were also included. These time windows were based on the timing of presentation (all within one calendar day of TDRS dilation) and treatment (all within 14 days of TDRS dilation) of all AAC cases identified by ICD codes.

**Table 1.** Demographic characteristics of the Los Angeles County Teleretinal Diabetic Retinopathy Screening.

| **Variable** |  | **Total population**<br>**n = 84,008** |
| --- | --- | --- |
| **Age at first dilation, mean (SD)** |  | 55.4 (10.7) |
| **Sex, n (%)** |  |  |
|  | Female | 46,255 (55.1) |
|  | Male | 36,074 (42.9) |
|  | Unknown | 1,679 (2.0) |
| **Race/ethnicity, n (%)** |  |  |
|  | White | 3,414 (4.1) |
|  | Black | 6,871 (8.2) |
|  | Asian | 5,301 (6.3) |
|  | Hispanic | 56,891 (67.7) |
|  | Unknown | 9,537 (11.4) |
|  | Other | 1,994 (2.4) |
| **Years with diabetes, n (%)** |  |  |
|  | 1 year or less | 14,469 (17.2) |
|  | 2-5 years | 23,897 (28.4) |
|  | 6-10 years | 18,449 (22.0) |
|  | 11-20 years | 17,347 (20.6) |
|  | >20 years | 8,932 (10.6) |
|  | Borderline | 779 (0.9) |
|  | Gestational | 47 (0.1) |
|  | Not diabetic | 41 (0.0) |
|  | Unknown | 47 (0.1) |
| **TDRS dilation count, mean (SD)** |  | 2.01 (1.50) |
| **TDRS dilation cohort, n (%)** |  |  |
|  | 1 dilation | 45,253 (53.9) |
|  | 2 dilations | 17,517 (20.9) |
|  | 3 dilations | 9,063 (10.8) |
|  | 4+ dilations | 12,175 (14.5) |
| **Median income by zip code** |  |  |
| | <\$40k | 769 (0.9) |
| | \$40k-\$49k | 4525 (5.4) |
| | \$50k-\$59k | 19697 (23.4) |
| | \$60k-\$74k | 24921 (29.7) |
| | \$75k-\$99k | 18875 (22.5) |
| | \$100k+ | 5806 (6.9) |
|  | Unknown | 9415 (11.2) |

Abbreviations: TDRS = teleretinal diabetic retinopathy screening

Manual chart review was performed for all identified encounters within the specified time windows; structured and unstructured EHR data fields were reviewed to confirm whether the encounter was related to a dilation-induced AAC attack. Additional demographic and clinical characteristics for AAC cases, including angle closure status in the non-presenting eye, time to presentation, presenting signs and symptoms, and treatment/s received, were also abstracted from the EHR. Clinical angle closure status was classified as primary angle closure suspect (PACS), primary angle closure (PAC), primary angle closure glaucoma (PACG), and acute primary angle closure (APAC) based on EHR data and definitions by Foster et al.^22^

### Statistical Analysis

Continuous data were expressed as means and standard deviations, and categorical data were expressed as proportions and percentages. The post-dilation AAC risk was estimated on both the dilation and patient levels. At the dilation level, AAC risk was calculated as total AAC events in at least one eye divided by total dilations. At the patient level, AAC risk was calculated as total patients who experienced an AAC event in at least one eye divided by total patients who underwent dilation. Statistical analysis was performed using R version 4.3.1.

## RESULTS

A total of 85,197 patients participated in 171,853 TDRS screening visits from August 2013 to March 2024. 63 patients were excluded due to missing or invalid identification numbers. An additional 1,126 patients deferred or did not receive dilation. The final cohort comprised 84,008 patients who underwent 168,796 total dilations (Figure 1).

**Figure 1.**
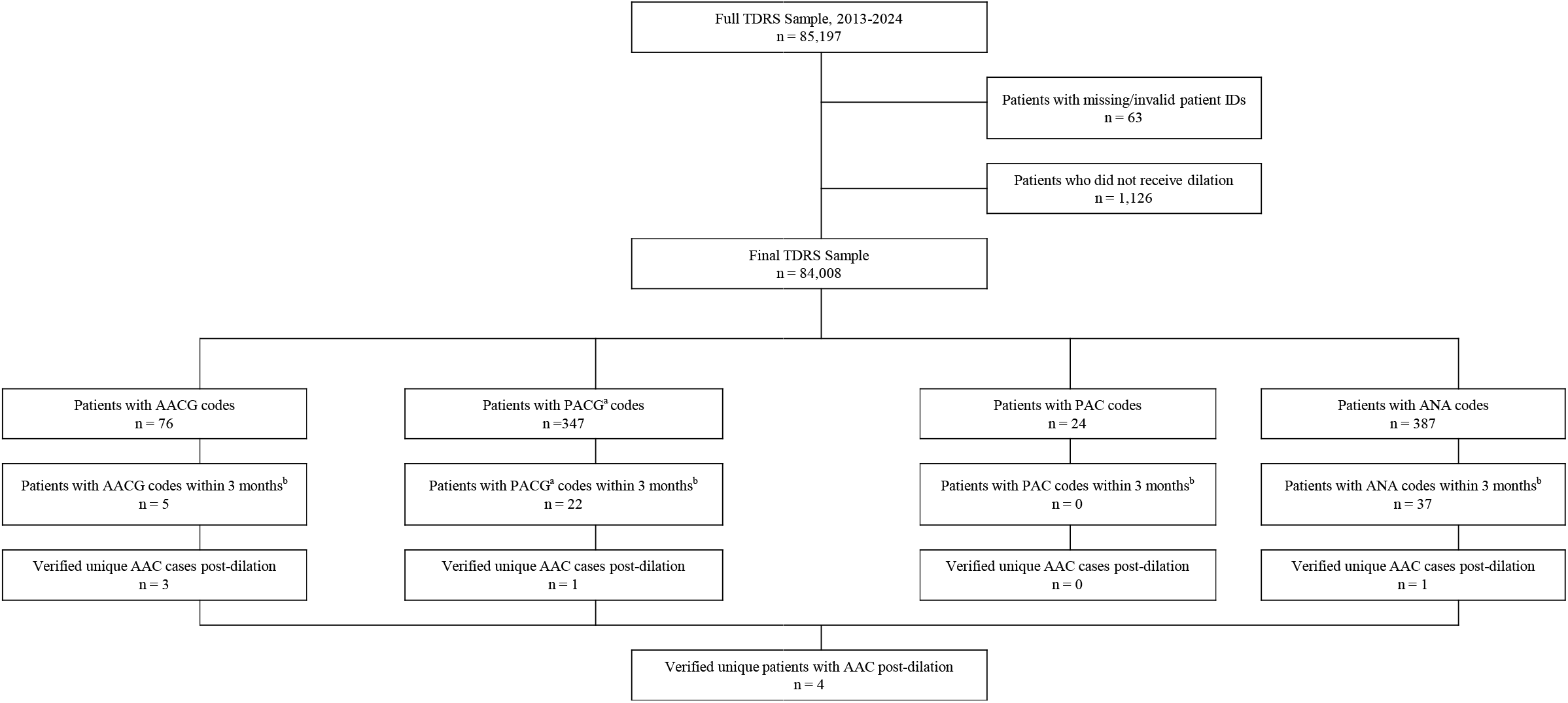
The diagram outlining the process of identifying patients with acute angle closure TDRS = teleretinal diabetic retinopathy screening; ID = identification; AACG = acute angle closure glaucoma; PACG = primary angle closure glaucoma; PAC = primary angle closure; ANA = anatomical narrow angle ^a^includes chronic, intermittent, residual, & unspecified PACG ^b^manual chart review was performed for these patients

Among these patients, 46,255 (55.1%) were female, 36,074 (42.9%) were male, and 1,679 (2.0%) were unspecified (Table 1). The mean age at first dilation was 55.4 ± 10.7 (mean ± standard deviation) years. 58.9% of the cohort was aged 60 years or less (See eFigure 1 in the Supplement). The cohort included 56,891 (67.7%) Hispanic, 6,871 (8.2%) Black, 5,301 (6.3%) Asian, 3,414 (4.1%) White, and 1,994 (2.4%) Other patients. The mean number of screening dilations was 2.01 ± 1.50 (mean ± standard deviation).

All encounters with ICD diagnosis codes for AACG (N=5), PACG (N=22), PAC (N=0), or ANA (N=37) within three months of a TDRS dilation were identified and underwent manual chart review. 60.0% (3/5 cases) of AACG, 4.5% (1/22 cases) of PACG, and 2.7% (1/37) of ANA cases were positive within 14 days of TDRS, yielding four AAC events in four unique patients after TDRS dilation (Figure 1). These four AAC events were associated with ICD codes for AACG (H40.21x), unspecified PACG (H40.20X*), and ANA (H40.03x). In one case, the AAC event was associated with overlap between an AACG (H40.21x) code and an unspecified PACG (H40.20X*) code. All four chart-confirmed AAC cases related to TDRS dilation presented acutely within one day of dilation. There were no additional AAC cases over the 3-month post-dilation period outside of the immediate 48-hour post-dilation window (See eFigure 2 in the Supplement). All cases received at least two ICD codes associated with the event, one in the UCC/ED at the initial encounter and another by an eye care provider at the subsequent referral visit (Table 3).

There were 157 patients with UCC/ED visits and 68 patients with eye clinic encounters who had eye-related diagnosis codes within one calendar day of TDRS dilation (Table 2). Seven additional patients received CPT codes for iridotomy/iridectomy or lens extraction within 14 calendar days of dilation. Manual chart review of these encounters identified all four AAC cases previously identified by ICD diagnosis codes but did not yield any additional cases. Of the 157 patients presenting to the UCC/ED, 113 had eye-related complaints outside of TDRS, 38 patients were referred by or advised at their TDRS visit to seek an UCC/ED evaluation for abnormalities identified on fundus photography or for more chronic visual complaints, and six patients had acute eye-related complaints following TDRS, including AAC, nausea, and subconjunctival hemorrhage. Of the 68 patients who presented to the eye clinic, 41 were referred at their TDRS visit or by the UCC/ED due to abnormalities identified on fundus photography or for more chronic visual complaints. Other eye clinic encounters occurring on the day of or within one day of TDRS were associated with follow-up/routine exams (n=23) or acute visual complaints unrelated to TDRS (n=4). Most patients (n=5) with CPT codes for iridotomy, iridectomy, or lens extraction received these treatments at routinely scheduled appointments unrelated to AAC, with two patients requiring iridotomy for urgent treatment of AAC.

**Table 2.** Patients with eye-related diagnoses presenting within one day after dilation.

| Location | Days Elapsed |  | Total |
| --- | --- | --- | --- |
|  | 0 | 1 |  |
| UCC/ED | 124 (0 AAC) | 33 (4 AAC) | 157 (4 AAC) |
| Eye Clinic | 41 (0 AAC) | 27 (0 AAC) | 68 (0 AAC) |

Abbreviations: UCC = Urgent Care Center; ED = Emergency Department; AAC = Acute Angle Closure

**Table 3.**
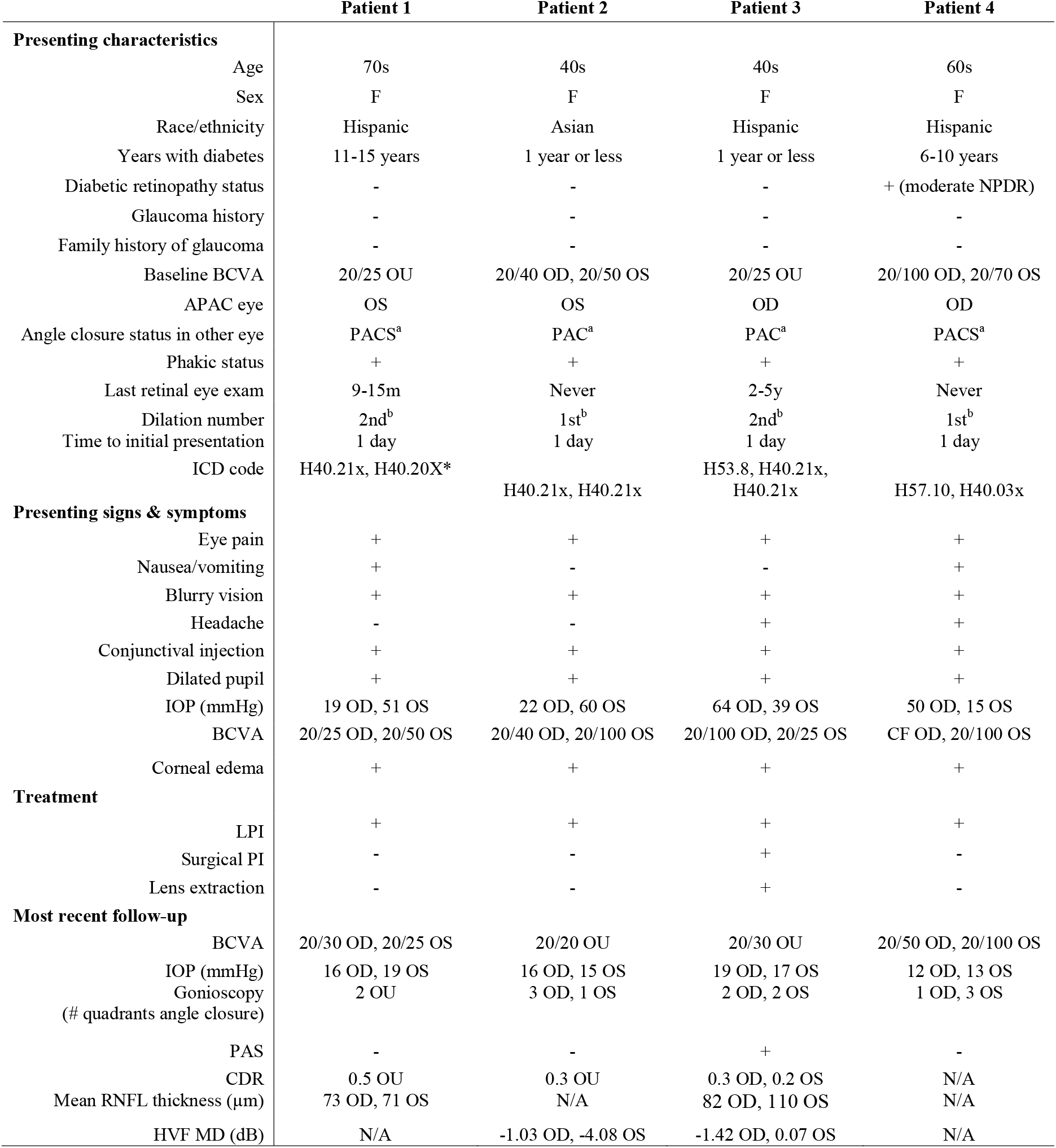
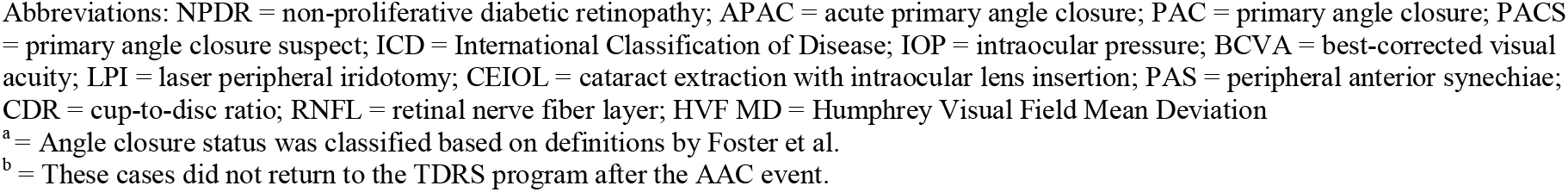
Demographic and clinical characteristics of the acute angle closure cases.

|  | Patient 1 | Patient 2 | Patient 3 | Patient 4 |
| --- | --- | --- | --- | --- |
| **Presenting characteristics** |  |  |  |  |
| Age | 70s | 40s | 40s | 60s |
| Sex | F | F | F | F |
| Race/ethnicity | Hispanic | Asian | Hispanic | Hispanic |
| Years with diabetes | 11-15 years | 1 year or less | 1 year or less | 6-10 years |
| Diabetic retinopathy status | - | - | - | + (moderate NPDR) |
| Glaucoma history | - | - | - | - |
| Family history of glaucoma | - | - | - | - |
| Baseline BCVA | 20/25 OU | 20/40 OD, 20/50 OS | 20/25 OU | 20/100 OD, 20/70 OS |
| APAC eye | OS | OS | OD | OD |
| Angle closure status in other eye | PACS<sup>a</sup> | PAC<sup>a</sup> | PAC<sup>a</sup> | PACS<sup>a</sup> |
| Phakic status | + | + | + | + |
| Last retinal eye exam | 9-15m | Never | 2-5y | Never |
| Dilation number | 2nd<sup>b</sup> | 1st<sup>b</sup> | 2nd<sup>b</sup> | 1st<sup>b</sup> |
| Time to initial presentation | 1 day | 1 day | 1 day | 1 day |
| ICD code | H40.21x, H40.20X* | H40.21x, H40.21x | H53.8, H40.21x,<br>H40.21x | H57.10, H40.03x |
| **Presenting signs & symptoms** |  |  |  |  |
| Eye pain | + | + | + | + |
| Nausea/vomiting | + | - | - | + |
| Blurry vision | + | + | + | + |
| Headache | - | - | + | + |
| Conjunctival injection | + | + | + | + |
| Dilated pupil | + | + | + | + |
| IOP (mmHg) | 19 OD, 51 OS | 22 OD, 60 OS | 64 OD, 39 OS | 50 OD, 15 OS |
| BCVA | 20/25 OD, 20/50 OS | 20/40 OD, 20/100 OS | 20/100 OD, 20/25 OS | CF OD, 20/100 OS |
| Corneal edema | + | + | + | + |
| **Treatment** |  |  |  |  |
| LPI | + | + | + | + |
| Surgical PI | - | - | + | - |
| Lens extraction | - | - | + | - |
| **Most recent follow-up** |  |  |  |  |
| BCVA | 20/30 OD, 20/25 OS | 20/20 OU | 20/30 OU | 20/50 OD, 20/100 OS |
| IOP (mmHg) | 16 OD, 19 OS | 16 OD, 15 OS | 19 OD, 17 OS | 12 OD, 13 OS |
| Gonioscopy<br>(# quadrants angle closure) | 2 OU | 3 OD, 1 OS | 2 OD, 2 OS | 1 OD, 3 OS |
| PAS | - | - | + | - |
| CDR | 0.5 OU | 0.3 OU | 0.3 OD, 0.2 OS | N/A |
| Mean RNFL thickness (µm) | 73 OD, 71 OS | N/A | 82 OD, 110 OS | N/A |
| HVF MD (dB) | N/A | -1.03 OD, -4.08 OS | -1.42 OD, 0.07 OS | N/A |

Abbreviations: NPDR = non-proliferative diabetic retinopathy; APAC = acute primary angle closure; PAC = primary angle closure; PACS = primary angle closure suspect; ICD = International Classification of Disease; IOP = intraocular pressure; BCVA = best-corrected visual acuity; LPI = laser peripheral iridotomy; CEIOL = cataract extraction with intraocular lens insertion; PAS = peripheral anterior synechiae; CDR = cup-to-disc ratio; RNFL = retinal nerve fiber layer; HVF MD = Humphrey Visual Field Mean Deviation
<sup>a</sup> = Angle closure status was classified based on definitions by Foster et al.
<sup>b</sup> = These cases did not return to the TDRS program after the AAC event.

Based on the number of verified AAC events, dilations, and patients dilated, the risk of post-dilation AAC was 2.4 (95% CI 0.05-4.69) per 100,000 (0.0024%) dilations in at least one eye, or 1 in 42,000 eyes dilated. The risk of post-dilation AAC was 4.8 (95% CI 0.10-9.43) per 100,000 (0.0048%) patients, or 1 in 21,000 patients dilated. The four AAC cases that occurred after TDRS dilation are described below (Table 3).

### Patient 1

The patient was a phakic Hispanic female in her 70s who presented to the ED one day following TDRS dilation complaining of unilateral vision loss OS. Best-corrected visual acuity (BCVA) was 20/25 OD and 20/50 OS. Intraocular pressure (IOP) was 19 mmHg OD and 51 mmHg OS. IV acetazolamide 500 mg was administered in the ED. Ophthalmology was consulted, and gonioscopy confirmed PACS OD and APAC OS. Bilateral laser peripheral iridotomies (LPIs) were performed. At the most recent follow-up, five months after presentation, BCVA was 20/30 OD and 20/25 OS with IOP of 16 mmHg OD and 19 mmHg OS on no medications.

### Patient 2

The patient was a phakic Asian female in her 40s who presented to the ED one day following TDRS dilation complaining of unilateral vision loss OS. BCVA was 20/40 OD and 20/100 OS. IOP was 22 mmHg OD and >60 mmHg OS. Oral acetazolamide 500 mg and IOP-lowering drops were administered in the ED. Ophthalmology was consulted, and gonioscopy confirmed PAC OD and APAC OS. Bilateral LPIs were performed. At the most recent follow-up, 54 months after presentation, BCVA was 20/20 OU with IOP of 16 mmHg OD and 15 mmHg OS on no medications.

### Patient 3

The patient was a phakic Hispanic female in her 40s who presented to the UCC one day following TDRS dilation with unilateral vision loss and conjunctival injection OD. The patient was managed conservatively for conjunctivitis and discharged home. She then presented to the ED one day later with persistent symptoms. BCVA was 20/100 OD and 20/25 OS. IOP was 64 mmHg OD and 39 mmHg OS. IV acetazolamide 500 mg and IOP-lowering drops were administered in the ED. Ophthalmology was consulted, and gonioscopy confirmed APAC OD and PAC OS. Bilateral LPI was attempted, with success OS and failure OD requiring urgent surgical iridectomy. The patient subsequently received lens extraction OU. At the most recent follow-up, one year after presentation, BCVA was 20/30 OU with IOP of 19 OD and 17 mmHg OS on 2 IOP-lowering drops.

### Patient 4

The patient was a phakic Hispanic female in her 60s who presented to the ED one day following TDRS dilation with unilateral vision loss. BCVA was CF OD and 20/100 OS. IOP was 50 mmHg OD and 15 mmHg OS. IV acetazolamide 500 mg and IOP-lowering drops were administered in the ED. Ophthalmology was consulted, and gonioscopy confirmed APAC OD and PACS OS. Right LPI was performed. At the most recent follow-up, one month after presentation, BCVA was 20/50 OD and 20/100 OS with IOP of 12 mmHg OD and 13 mmHg OS on 4 IOP-lowering drops OD.

## DISCUSSION

We found a very low incidence of AAC after pharmacologic dilation in the LAC DHS TDRS population, with only four identified AAC cases from 168,796 dilated screening exams. All were diagnosed by the next full calendar day and received LPI within one week of dilation. Manual chart review confirmed that the ICD code for AACG (H40.21x) was highly specific for identifying post-dilation AAC, even in a capitated payment system not reliant on accurate coding for reimbursement. These findings support the safety of mydriatic fundus photography in teleretinal screening programs and the use of ICD codes in claims-based studies on AAC.

The incidence of AAC was 2.4 per 100,000 dilations (0.0024%) or less than 1 in 40,000 dilations, closely aligning with other teleretinal screening programs. For example, the Northern Ireland Diabetic Screening Program (NIDRSP) reported an AAC risk of 1 in 31,755 dilations.^12^ The exceedingly low incidence in both studies underscores the safety of dilations in large-scale eye disease screening programs, even among diabetic patients. In the LAC DHS TDRS program, less than 1% (583 out of 187,963) of fundus photographs are ungradable (unpublished data). Therefore, it is imperative to continue the discussion on how to balance the benefits of dilation for earlier and more accurate detection of eye disease against the very low risk of AAC after dilation.

It is also important to contextualize our post-dilation AAC risk estimates with previous studies on PACG prevalence and AAC rates after pharmacologic dilation in angle closure eyes. The Zhongshan Angle-Closure Prevention (ZAP) Study, a study of mainland Chinese primary angle closure suspects (PACS), reported a relatively high AAC incidence of 0.06%, which is 25 times higher than our AAC risk estimate (0.0024%).^8^ Therefore, the prevalence of angle closure would need to be around 4.0% or lower to be consistent with the low rate of AAC in our study. Given the very low prevalence of PACG (0.1%) reported in Proyecto Ver, another study of predominantly Hispanic eyes, the prevalence of angle closure in our study cohort was likely even lower than 4.0%. In addition, the mean age of our cohort was younger (60.3 ± 13.2 years) than Proyecto Ver (76.0 ± 9.2 years), further skewing our cohort away from angle closure at baseline.^23^ For further context, the Baltimore Study, which excluded patients with narrow angles, reported no cases of AAC after dilation in 4,870 White or Black participants.^7^ These data alleviate concerns about severe underdetection of AAC and increase the plausibility of our post-dilation AAC risk estimates, which may initially seem low in the context of previous studies on angle closure eyes.

We constructed and validated an AAC phenotype by cross-referencing ICD codes with EHR data and conducting a manual chart review. Three of the four verified AAC cases received an AACG code within one day of dilation. The fourth verified AAC case presented to the ED within one day of TDRS dilation but did not receive an AACG code. This case led us to broaden our search strategy and perform a manual chart review of all acute presentations to the ED, UCC, or eye clinic with eye diagnosis codes within one full calendar day after TDRS and all visits with procedure codes within 14 days of TDRS, as each AAC case received LPI within one week of TDRS dilation. There were no additional miscoded/misdiagnosed AAC cases, validating our initial strategy for identifying post-dilation AAC cases. All cases were associated with at least two separate encounters, at least one of which involved an eye care professional. Future investigations using claims data, particularly when EHR access is unavailable, may improve diagnostic accuracy by incorporating a short time window from dilation to AACG diagnosis code, an eye care visit for phenotype confirmation, and an additional LPI treatment code.

Although our sample size was insufficient to model risk factors for post-dilation AAC, the four AAC cases shared commonalities in their presentation and outcomes. All were female, phakic, had narrow angles in the non-presenting eye identified on gonioscopy, and presented within one day with AAC symptoms, including eye pain and blurry vision. All AAC attacks occurred in phakic eyes, highlighting the crucial role the lens plays in pupillary block and angle closure mechanisms.^24^ Interestingly, two of the four cases were under 50 years old; this finding appears to implicate non-lens related mechanisms (e.g. plateau iris, thick iris roll) over lens-related (e.g. pupillary block, high lens vault) mechanisms in the pathogenesis of AAC, which remains poorly understood.^25^ In addition, one case occurred in an Asian patient, a group that comprised of only 6.3% of the cohort. This finding (1 post-dilation AAC case in approximately 530 Asians with angle closure assuming a 10% prevalence of angle closure) approximates findings by Lavanya et al. and implicates Asian race as a potential risk factor for post-dilation AAC.^8-11^ Finally, all TDRS patients received detailed AAC return precautions at time of dilation, which may explain why all four cases presented shortly after AAC onset, received prompt ophthalmic care, and had good visual outcomes despite potential barriers to care in this under-resourced safety net health system. While Patient 4 had relatively poor follow-up BCVA, we note that her visual acuity was already reduced at the initial TDRS visit, likely due to underlying diabetic retinopathy. These outcomes highlight the importance of patient education at time of screening and overall safety of the LAC DHS TDRS program even when complicated by a rare AAC attack.

Our study assessed AAC risk in a unique patient population, as LAC DHS primarily serves socioeconomically disadvantaged racial minorities who typically encounter more barriers to care. Most of our cohort resided in zip codes with a median household income of $74,000 or less, which was below the 2022 median household income of $83,411 for Los Angeles County.^26^ Patients of lower socioeconomic status are less likely to seek or receive timely medical attention or treatment.^27^ Furthermore, our study population was predominantly Hispanic. Hispanic ethnicity has been associated with lower health literacy, potentially contributing to greater avoidance of healthcare services, including follow-up ophthalmological care.^27-31^ These characteristics of our safety net population may contribute to our low incidence of AAC, with individuals less likely to seek or receive care compared to the general population. Finally, our findings highlight the importance of continued access to eye care after TDRS screening, which may be challenging for patients living in remote or less developed regions.

Our study has some limitations. First, it can be difficult to determine whether an AAC diagnosis resulted from dilation relying solely on claims data. We were able to mitigate this issue by cross-referencing ICD codes with patient EHR data and conducting a manual chart review. Second, some patients experiencing AAC may have sought care outside of the LAC DHS system despite being instructed to visit an LAC DHS UCC/ED in case of angle closure symptoms. However, given that TDRS patients receive all their regular healthcare within the LAC DHS system, and every TDRS facility has an ED and/or UCC, it is likely that LAC DHS patients would seek emergent/urgent care at an LAC DHS facility. Third, patients with more severe angle closure (e.g. PACG or prior episode of AAC) may have been previously detected and established with in-office care, skewing the TDRS population toward healthier patients at lower risk for AAC. However, this bias exists in all teleretinal screening programs, which tend to monitor lower risk patients and refer higher risk patients for in-office care.

In conclusion, we identified a very low risk of AAC following pharmacologic dilation in a large safety net TDRS population, reinforcing the overall safety of such screening programs and practices. We also validated the use of ICD codes to identify AAC cases, supporting future research on AAC using other databases, such as nationwide EHR or healthcare claims databases. Further discussion about the safety of pharmacologic dilation is crucial for optimizing teleretinal screening workflows, especially with the development of artificial intelligence (AI) algorithms and data tools that rely on high-quality mydriatic fundus photos.^32-33^ Future research should also aim to identify factors conferring greater risk of AAC and developing convenient diagnostic tools, possibly using OCT imaging, to identify individuals in whom pharmacologic dilation should be avoided.^34-38^

## Data Availability

All data produced in the present work are contained in the manuscript.

