## Supplemental Table 1 for "Safety of Pharmacologic Dilation: Acute Angle Closure Incidence in a Los Angeles County-Wide Safety Net Teleretinal Screening Program"

**eTable 1. Current Procedural Terminology Codes for Iridotomy, Iridectomy, and Lens Extraction**

Iridotomy/iridectomy:

| 66761 | Iridotomy/iridectomy by laser surgery |
| --- | --- |
| 66840 | Removal of lens material; aspiration technique, 1 or more stages |
| 66762 | Iridoplasty by photocoagulation |
| 65820 | Iridotomy, laser, anterior chamber |
| 65830 | Iridectomy, anterior chamber, with or without peripheral iridectomy |
| 66625 | Peripheral iridectomy with a corneoscleral or corneal section for glaucoma |
| 66630 | Iridectomy with a corneoscleral or corneal section for glaucoma in a sector |
| 66500 | Iridotomy by stab incision (separate procedure); except transfixion |
| 66505 | Iridotomy by stab incision (separate procedure) with transfixion as for iris bombe |

Lens extraction:

| 66982 | Complex extracapsular cataract removal with IOL insertion, requiring specialized techniques or devices |
| --- | --- |
| 66984 | Extracapsular cataract removal with insertion of intraocular lens prosthesis (1 stage procedure), manual or mechanical technique; without endoscopic cyclophotocoagulation |
| 66987 | Extracapsular cataract removal with insertion of intraocular lens prosthesis (1 stage procedure), manual or mechanical technique, complex; with endoscopic cyclophotocoagulation |
| 66988 | Extracapsular cataract removal with insertion of intraocular lens prosthesis (1 stage procedure), manual or mechanical technique; with endoscopic cyclophotocoagulation |
| 66940 | Extracapsular lens material removal |
