## Supplementary figures and images for "Safety of Pharmacologic Dilation: Acute Angle Closure Incidence in a Los Angeles County-Wide Safety Net Teleretinal Screening Program"

### Supplemental Figure 1

**eFigure 1. Histogram of Age Distribution**

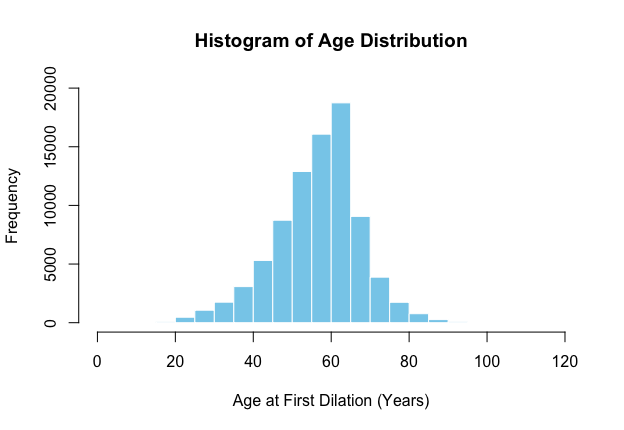

### Supplemental Figure 2

**eFigure 2. Histogram of Time to Confirmed AAC Presentation**

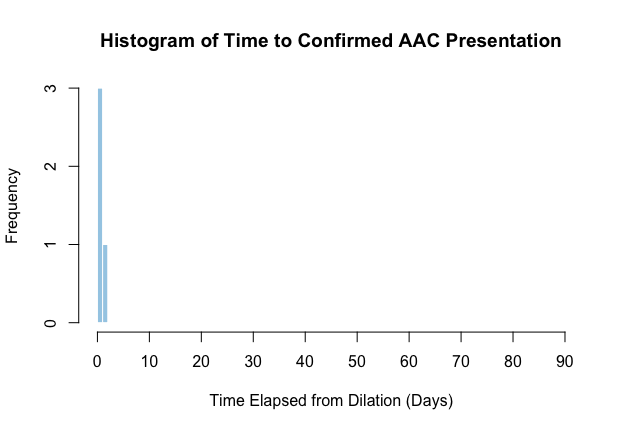
